# Critical Role for 24-Hydroxylation in Homeostatic Regulation of Vitamin D Metabolism

**DOI:** 10.1101/2023.06.27.23291942

**Authors:** Zhinous Shahidzadeh Yazdi, Elizabeth A. Streeten, Hilary B. Whitlatch, May E. Montasser, Amber L. Beitelshees, Simeon I. Taylor

## Abstract

**Context:** The body has evolved homeostatic mechanisms to maintain free levels of Ca^+2^ and 1,25-dihydroxyvitamin D [1,25(OH)_2_D] within narrow physiological ranges. Clinical guidelines emphasize important contributions of PTH in maintaining this homeostasis.

**Objective:** To investigate mechanisms of homeostatic regulation of vitamin D (VitD) metabolism and to apply mechanistic insights to improve clinical assessment of VitD status.

**Design:** Crossover clinical trial studying participants before and after VitD3-supplementation.

**Setting:** Community.

**Participants:** 11 otherwise healthy individuals with VitD-deficiency (25-hydroxyvitamin D [25(OH)D] ≤20 ng/mL).

**Interventions:** VitD3-supplements (50,000 IU once or twice a week depending on BMI, for 4-6 weeks) were administered to achieve 25(OH)D≥30 ng/mL.

**Results:** VitD3-supplementation significantly increased mean 25(OH)D by 2.7-fold and 24,25-dihydroxyvitamin D [24,25(OH)_2_D] by 4.3-fold. In contrast, mean levels of PTH, FGF23, and 1,25(OH)_2_D did not change. Mathematical modeling suggested that 24-hydroxylase activity was maximal for 25(OH)D≥50 ng/mL and achieved a minimum (∼90% suppression) with 25(OH)D<10-20 ng/mL. The 1,25(OH)_2_D/24,25(OH)_2_D ratio better predicted modeled 24-hydroxylase activity (*h*) (ρ=-0.85; p=0.001) compared to total plasma 25(OH)D (ρ=0.51; p=0.01) and the 24,25(OH)_2_D/25(OH)D ratio (ρ=0.37; p=0.3).

**Conclusions:** Suppression of 24-hydroxylase provides a first line of defense against symptomatic VitD-deficiency by decreasing metabolic clearance of 1,25(OH)_2_D. The 1,25(OH)_2_D/24,25(OH)_2_D ratio provides a useful index of VitD status since it incorporates 24,25(OH)_2_D levels and therefore, provides insight into 24-hydroxylase activity. When VitD availability is limited, this suppresses 24-hydroxylase activity – thereby decreasing the level of 24,25(OH)_2_D and increasing the 1,25(OH)_2_D/24,25(OH)_2_D ratio. Thus, an increased 1,25(OH)_2_D/24,25(OH)_2_D ratio signifies triggering of homeostatic regulation, which occurs at early stages of VitD-deficiency.

## INTRODUCTION

Vitamin D (VitD) plays an essential role in mediating absorption of dietary calcium in the intestine (1–4). Furthermore, as evidenced by diseases such as nutritional rickets, VitD is critical to maintaining bone health (5, 6). The body obtains VitD from two sources. Endogenous production of VitD requires exposure to ultraviolet light, which is variable depending on geographic location and the season of the year among other factors. Dietary intake of VitD is also variable and depends on the VitD content of the diet (including intake of fortified dairy products and dietary supplements) (7, 8). To maintain a consistent *milieu intérieure* (9, 10), each individual must maintain levels of free 1,25-dihydroxyvitamin D [1,25(OH)_2_D] – the biologically active metabolite of VitD – within a narrow physiological range notwithstanding considerable variation in availability of the biosynthetic precursor (i.e., VitD) (11–13).

We developed a mathematical model to investigate physiological mechanisms mediating homeostatic regulation of VitD metabolism. 24-hydroxylation provides the major mechanism whereby 1,25(OH)_2_D is cleared (12, 14, 15). According to our mathematical model, *in vivo* activity of 24-hydroxylase varies over an approximately tenfold range. 24-Hydroxylase activity achieves a maximum when 25(OH)D levels exceed 50 ng/mL. By inducing 24-hydroxylase activity when VitD is abundant, the body increases clearance of 1,25(OH)_2_D – thereby protecting against VitD toxicity. Furthermore, the model suggests that 24-hydroxylase activity achieves a minimum when 25(OH)D is less than 10-20 ng/mL. The resultant decrease in 1,25(OH)_2_D clearance tends to sustain 1,25(OH)_2_D levels, thereby compensating for limited availability of VitD and protecting against symptomatic VitD deficiency.

Several feedback loops contribute to homeostatic regulation of VitD – including feedback loops between VitD status and the rates of both production and clearance of 1,25(OH)_2_D (12, 16–19). Cannon proposed the concept of the “wisdom of the body” (9), which includes homeostatic mechanisms that are responsible for maintaining a constant *milieu intérieure.* Endocrinologists often leverage the “wisdom of the body” to diagnose states of hormonal excess or hormonal insufficiency based on the level of activation or suppression of homeostatic mechanisms. This is exemplified by the approach to diagnosing patients’ hormonal status with respect to hormones that act upon other nuclear hormone receptors (e.g., thyroid hormones or glucocorticoids) by measuring the levels of trophic hormones (e.g., TSH or ACTH) (20, 21). By analogy, we discuss our data in the context of the published literature advocating for a similar approach to assessing VitD status – i.e., by assessing the level of suppression/induction of 24-hydroxylase activity (22–24). By leveraging the wisdom of the body in this assessment, physicians can advance the state-of-the-art of assessment of VitD status.

The present work is a supplemental analysis of data obtained in a clinical trial investigating the nutrient-drug interaction between VitD and canagliflozin (13). That clinical trial provided data on the responses of PTH and VitD metabolites to VitD3 supplementation in participants whose baseline 25(OH)D was in the range of 10-20 ng/mL. We developed a mathematical model to investigate regulatory mechanism mediating these responses and also considered the clinical implications of the predictions by this model.

## METHODS

This study (“VitD Sub-study”) was part of our ongoing “Genetics of Response to Canagliflozin (GRC)” clinical trial (Clinicaltrials.gov Identifier: NCT02891954) in healthy volunteers recruited from the Old Order Amish population in Lancaster PA, who are non-Hispanic white people. Although participants were excluded from the parent study if their 25(OH)D level ≤20 ng/mL, these VitD deficient individuals were eligible to participate in the VitD Sub-study, which investigated the effects of VitD3 supplements on canagliflozin-triggered changes in the serum phosphorus/FGF23/1,25(OH)_2_D/PTH axis (13). Participants were required to meet all other inclusion/exclusion criteria for the parent study. The effect of VitD3 supplementation on canagliflozin-induced changes in mineral metabolism biomarkers was reported elsewhere (13). The current paper reports results of a secondary data analysis focusing primarily on the effect VitD3 supplementation on VitD metabolites in participants with mild-moderate VitD deficiency.

### Screening and recruitment

As reported previously, we screened 24 healthy volunteers from September 2019 to July 2021 and enrolled 19 individuals with 25(OH)D levels ≤20 ng/mL who were otherwise eligible to participate in the parent GRC study (13). The Endocrine Society’s clinical practice guidelines proposed thresholds of 25(OH)D ≤20 ng/mL (≤50 nmol/L) for VitD deficiency, 21-29 ng/mL (51-74 nmol/L) for VitD insufficiency, and ≥ 30 ng/mL (≥75 nmol/L) for VitD sufficiency (17). We applied The Endocrine Society’s definition of VitD deficiency (≤20 ng/mL) as a key inclusion criterion for this sub-study and applied the definition of VitD sufficiency (≥30 ng/mL) as the criterion for adequacy of VitD3 supplementation. Details of demographics, baseline characteristics, recruitment, and disposition are reported elsewhere (13). Because of interruptions and delays associated with the COVID-19 pandemic, we did not complete our plan to study 25 research participants. We analyzed data after eleven participants (four females and seven males) completed the entire study protocol.

### Endpoints and biomarkers

We measured levels of the following biomarkers both at baseline and after VitD3 supplementation: plasma PTH, plasma FGF23, serum phosphorus, serum calcium, and three VitD metabolites – 25(OH)D, 1,25(OH)_2_D, and 24,25-dihydroxyvitamin D [24,25(OH)_2_D].

### VitD3 supplementation protocol

VitD deficient participants completed canagliflozin challenge #1 and then received VitD3 capsules (Bio-Tech Pharmacal; Fayetteville, AR) in doses of either 50,000 IU per week for participants with BMI <30 kg/m^2^ or 50,000 IU twice a week for participants with BMI ≥30 kg/m for four weeks, followed by a second total serum 25(OH)D measurement a week after completion of the four-week VitD3 supplementation protocol (13). If screening serum 25(OH)D levels (Quest Diagnostics) were <30 ng/mL, high-dose VitD3 supplementation was continued. Once screening 25(OH)D (Quest Diagnostics) levels were confirmed to be ≥30 ng/mL, participants were administered the maintenance dose of VitD3 supplementation (1000 IU/d or 2000 IU/d for BMI <30 kg/m^2^ or BMI ≥30 kg/m^2^, respectively) for at least six weeks to allow them to re-equilibrate to a lower physiologic dose and to develop a new steady state. The maintenance dose was continued until completion of canagliflozin challenge #2.

### Clinical chemistry

Blood samples obtained at home visits were collected in test tubes as appropriate for each assay (13). Quest Diagnostics conducted an immunoassay for baseline levels of plasma PTH as well as LC-MS/MS assays to measure baseline serum 1,25(OH)_2_D, and serum 25(OH)D levels. Plasma levels of 25(OH)D and 24,25(OH)_2_D were assessed using LC-MS/MS assays conducted by Heartland Assays (Ames, IA). Quest’s serum 25(OH)D levels were used to determine eligibility for the VitD Sub-study. Heartland’s plasma 25(OH)D levels were used to assess the response to VitD3 supplements. Participants were recruited to the parent study, which used Quest’s 25(OH)D assay to screen for eligibility. Therefore, eligibility for the sub-study was also based on Quest’s assay. The VitD sub-study used Heartland’s assay to obtain data on 24,25(OH)2D levels. We also used Heartland’s 25(OH)D assay to obtain data on 25(OH)D levels on the same day we obtained all the other data in the study. In contrast, the screening data on 25(OH)D were typically obtained at least one to two weeks prior to conducting the actual studies. We wanted to compare 25(OH)D levels on the day of screening to the 25(OH)D level on the day of the actual study to determine whether 25(OH)D levels had changed during that time interval. We have reported 25(OH)D and 24,25(OH)_2_D in ng/mL and 1,25(OH)_2_D in pg/mL. Multiplication by a factor of 2.49 can be used to convert from ng/mL to nmol/L. Mean levels for 25(OH)D before and after VitD3 supplementation, were identical in assays conducted by Quest Diagnostics and Heartland Assays (13). LC-MS/MS assays conducted at both Quest Diagnostics and Heartland Assays distinguished between derivatives of VitD2 and VitD3. Derivatives of VitD2 were not detected in any of our participants’ samples. For simplicity, we refer to our data on levels of 25(OH)D, 1,25(OH)_2_D, or 24,25(OH)_2_D without specifying D2 or D3. Nevertheless, since D2 derivatives were not detected in any of our participants, this should be understood as referring exclusively to D3 derivatives.

### Quality control for VitD assays of VitD metabolites

Heartland Assays provided the following information regarding their assays of 25(OH)D and 24,25(OH)_2_D: (a) accuracy >90% based on comparisons to National Institute of Standards and Technology standards; and (b) coefficients of variation for both inter- and intra-assay comparisons were < 5%.

### Analysis of our data

We obtained blood samples on three separate days during canagliflozin (300 mg/day) challenge tests: baseline prior to receiving canagliflozin, at 48 hours (after receiving canagliflozin for two days), and at 120 hours (after receiving canagliflozin for five days). Because mean levels of 25(OH)D and 24,25(OH)_2_D were not affected by canagliflozin, we calculated coefficients of variation based on an assumption that the three separate blood samples obtained from each individual during the canagliflozin challenge test had similar concentrations of those two VitD metabolites. Based on this assumption, we calculated coefficients of variation of 5.8% for 25(OH)D and 11.1% for 24,25(OH)_2_D. Because these calculated coefficients of variation included day-to-day variation in VitD metabolite levels as well as intra-assay variation, our estimates of coefficients of variation are likely greater than the actual intra-assay variation.

Furthermore, we compared mean data for 25(OH)D levels measured in the same individuals in the two assays. Mean 25(OH)D levels before receiving VitD3 supplements were 16.4 and 16.5 ng/mL in the Quest and Heartland assays, respectively (13). Mean total 25(OH)D levels after receiving VitD3 supplements were 45.3 and 44.3 ng/mL in the Quest and Heartland assays, respectively (13). In addition, the individual data from the Quest assay (blood obtained on the day of screening) was highly correlated with individual data from the Heartland assay (blood obtained on Day 1 of the canagliflozin challenge test) – with variance in one assay accounting for 81% of the variance in the other assay. Based on these data, we conclude that the two assays provide comparable measurements of 25(OH)D levels. In any case, the Quest assay for 25(OH)D was used only to determine eligibility for the study. The Heartland assay was the sole source of data on 25(OH)D levels used for determination of goodness-of-fit for our model.

### Published analysis of Quest’s assay of 1,25(OH)_2_D

Biancuzzo et al. (25) reported intra- and inter-assay coefficients of variation of 9% and 12%, respectively, for Quest’s assay of 1,25(OH)_2_D. In addition, we estimated the inter-assay coefficient of variation based on our observation that 1,25(OH)_2_D levels were not changed by administration of VitD3 supplements. When we calculated a coefficient of variation for 1,25(OH)_2_D levels before and after VitD3 supplementation, we estimated a coefficient of variation of 13% – very similar to the inter-assay coefficient of variation reported by Biancuzzo et al. (25). Our calculation of coefficient of variation includes at least three components: the coefficient of variation for measurement error, day-to-day variation, and a possible small effect of VitD3 supplementation on 1,25(OH)_2_D levels. Thus, our estimate of the coefficient of variation represents an upper bound for the actual inter-assay coefficient of variation.

### Mathematical Modeling

We developed a mathematical model to analyze relationships among 25(OH)D, 1,25(OH)_2_D, and 24,25(OH)_2_D. See Table 1 for definitions of model parameters. For the purpose of fitting curves to model our clinical trial data, we estimated model parameters using an iterative trial and error approach.

**Table 1.** Parameters of the mathematical model.

| **Model parameters** | **Definitions** |
| --- | --- |
| **$\alpha$** | A model parameter to facilitate fitting the equation to the observed data |
| **$h$** | Relative 24-hydroxylase activity |
| **$K_I$** | Michaelis-Menten constant for 1α-hydroxylase with 25(OH)D as substrate |
| **$K_{24}$** | Michaelis-Menten constant for 24-hydroxylase enzyme with 25(OH)D as substrate (to fit hyperbolic curve) |
| **$L_{1,25}$** | Analog of Michaelis-Menten constant for 24-hydroxylase enzyme with 1,25(OH)<sub>2</sub>D as substrate (to fit sigmoid curve) |
| **$L_{24}$** | Analog of Michaelis-Menten constant for 24-hydroxylase enzyme with 25(OH)D as substrate (to fit sigmoid curve) |
| **$m(1,25)$** | First order rate constant for metabolic clearance of 1,25(OH)<sub>2</sub>D |
| **$m(24,25)$** | First order rate constant for metabolic clearance of 24,25(OH)<sub>2</sub>D |
| **$S(1,25)$** | 1,25(OH)<sub>2</sub>D concentration |
| **$S(24,25)$** | 24,25(OH)<sub>2</sub>D concentration |
| **$S(25)$** | 25(OH)D concentration |
| **$V_I$** | Maximal velocity for 1α-hydroxylase with 25(OH)D as substrate |
| **$V_{24}$** | Maximal velocity for 24-hydroxylase with 25(OH)D as substrate (to fit hyperbolic curve) |
| **$W_{1,25}$** | Maximal velocity for 24-hydroxylase with 1,25(OH)<sub>2</sub>D as substrate (to fit sigmoid curve) |
| **$W_{24}$** | Maximal velocity for 24-hydroxylase with 25(OH)D as substrate (to fit sigmoid curve) |

The observed sigmoid curve expressing the concentration of total 24,25(OH)_2_D as a function of total 25(OH)D was fitted by an equation with the form of Equation (1):

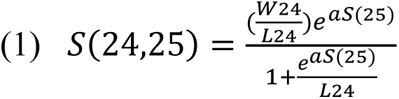

where S(25) and S(24,25) represent levels of 25(OH)D and 24,25(OH)_2_D, respectively; W_24_ and L_24_ represent analogs of the maximal velocity and Michaelis-Menten constant for 24-hydroxylase enzyme, respectively, with 25(OH)D as substrate; α is a model parameter to facilitate fitting the equation to the observed data.

The model assumes that 24,25(OH)_2_D is cleared by a first order process with a rate constant of *m*(*24,25*) [Equation (1a)]:

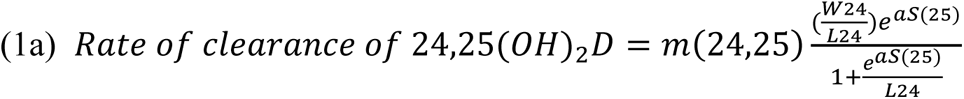

We hypothesized that the 24-hydroxylase enzyme follows Michaelis-Menten kinetics as described by Equation (2):

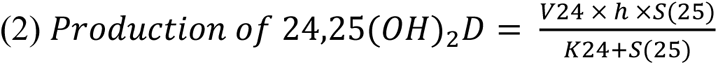

where S(25) represents the level of 25(OH)D; V_24_ and K_24_ represent the maximal velocity and Michaelis-Menten constant for 24-hydroxylase enzyme, respectively, with 25(OH)D as substrate. *h* is a variable function of S(25) that represents 24-hydroxylase activity as a fraction of the maximally induced level of the enzyme observed when 25(OH)D levels are high.

At steady state, the rate of production of 24,25(OH)_2_D [Equation (2)] is equal to the rate at which 24,25(OH)_2_D is cleared [Equation (1a)]. Accordingly, the value of *h* can be calculated by Equation (3):

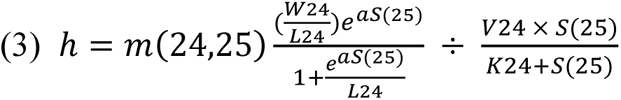

Genetic evidence demonstrates the critical role of CYP24A1-mediated 24-hydroxylation in inactivation of 1,25(OH)_2_D (26, 27). Homozygosity for loss of function mutations in *CYP24A1* has been reported to cause hypercalcemia – presumably due to excessive levels of 1,25(OH)_2_D. Since CYP24A1 catalyzes 24-hydroxylation of both 25(OH)D and 1,25(OH)_2_D, we modified Equation (1a) to yield Equation (4):

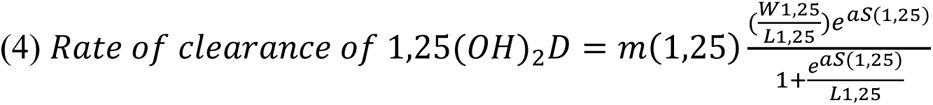

where *S(1,25)* represents the level of 1,25(OH)_2_D; *W_1,25_* and *L_1,25_* represent analogs of the maximal velocity and Michaelis-Menten constant for 24-hydroxylase enzyme, respectively, with 1,25(OH)_2_D as substrate; α is a model parameter that facilitates fitting the equation to the observed data; and *m(1,25)* is a first-order rate constant analogous to *m(24,25)* in Equation (1a). Further, we hypothesize that CYP27B1-mediated 1α-hydroxylation of 25(OH)D follows Michaelis-Menten kinetics:

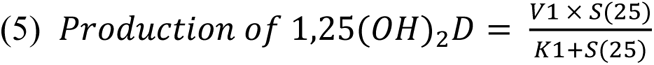

where V_1_ and K_1_ represent the maximal velocity and Michaelis-Menten constant for 1α-hydroxylase, respectively, with 25(OH)D as substrate. Under steady state conditions, the rates of production and metabolic clearance of 1,25(OH)_2_D are equal. Thus, we can combine Equations (4) and (5) to yield Equation (6):

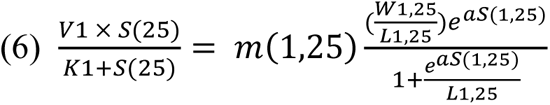

Solving for *S(1,25)*, we obtain Equation (7):

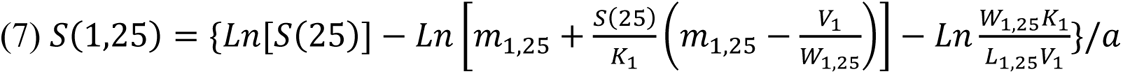

### Goodness of fit

We calculated the following metrics in order to evaluate the goodness of fit between our mathematical model and the experimental data:

Sum of squares due to regression: ***SSR*** = Σ(*x̂_i_* – *x̅*)^2^

Total sum of squares about the mean: ***SST*** = Σ(*x_i_* – *x̅*)^2^

Sum of squared errors: ***SSE*** = Σ(*x_i_* – *x̂*)^2^

Data on goodness of fit is provided in the Results section as appropriate.

### Estimating relative 24-hydroxylase activity

Equation (3) expresses relative 24-hydroxylase activity (*h*) as a function of the 25(OH)D level. However, total 25(OH)D levels are affected by inter-individual variation in VDBP levels, a factor which is not included in our model. Accordingly, we replaced 25(OH)D levels by the ratio of 25(OH)D/1,25(OH)_2_D. The 25(OH)D/1,25(OH)_2_D ratio is independent of VDBP level and provides an index of free levels of 25(OH)D. To normalize values of this ratio to correspond to values of total 25(OH)D, we first divided individual values of the 25(OH)D/1,25(OH)_2_D by a normalization factor [the mean value of the 25(OH)D/1,25(OH)_2_D ratio before administration of VitD3 supplements]. We next converted individual values of the normalized 25(OH)D/1,25(OH)_2_D ratio to the same units as total 25(OH)D by multiplying by a conversion factor [the mean value of 25(OH)D before administration of VitD3 supplements].

### Statistics

A two-sided p-value <0.05 (paired t-test) was taken as the threshold for statistical significance without any correction for multiple comparisons. We pooled all the data before and after VitD3 supplementation for graphing total levels of 1,25(OH)_2_D and 24,25(OH)_2_D as a function of total levels of 25(OH)D. Both Pearson and Spearman correlation coefficients were calculated using software provided in Excel.

### Study approval

The Clinical Trial Protocol was approved by the University of Maryland Baltimore Institutional Review Board (FWA00007145). Written informed consent was obtained from all participants.

## RESULTS

### Interrelationships among 1,25(OH)_2_D, 24,25(OH)_2_D, and 25(OH)D

As reported previously, mean (±SEM) total plasma levels of both 25(OH)D and 24,25(OH)_2_D increased significantly in response to VitD3 supplementation: from 16.5±1.6 to 44.3±5.5 ng/mL (p=0.0006) for 25(OH)D and from 1.0±0.1 to 4.3±0.6 ng/mL (p=0.0002) for 24,25(OH)_2_D (13). We observed consistent increases in total levels of 25(OH)D and 24,25(OH)_2_D (Figs. 1A and 1B). In contrast, mean total plasma levels of 1,25(OH)_2_D did not change in response to VitD3 supplements (43.8±3.6 pg/mL versus 44.9±4.1 pg/mL; p=0.7) (Fig. 1C) (13). This observation provides strong evidence of the effectiveness of homeostatic mechanisms regulating levels of 1,25(OH)_2_D. Therefore, we conducted exploratory analyses to further investigate physiological mechanisms mediating this homeostatic regulation.

**Figure 1.**
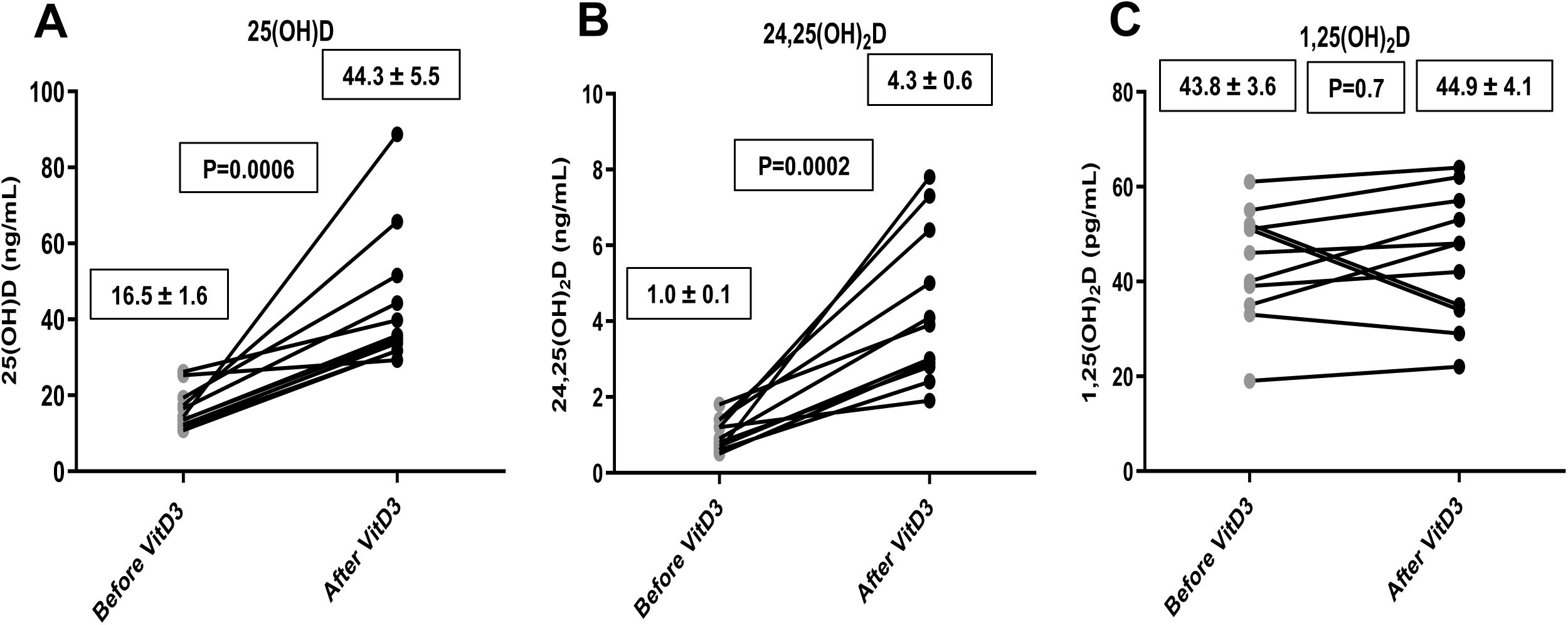
Impact of VitD3 supplementation on total levels of VitD metabolites. Healthy participants (N=11) were recruited based on meeting the Endocrine Society’s criterion of VitD deficiency [25(OH)D ≤20 ng/mL]. Total levels of VitD metabolites were assayed before and after participants received VitD3 supplements (13): 25(OH)D (panel A), 24,25(OH)_2_D (panel B), and 1,25(OH)_2_D (panel C). Data are presented as means ± SEM; p-values were calculated using a two-tailed t-test for paired data.

For this analysis, we pooled data on VitD metabolites – including data obtained both before and after VitD3 supplementation (Fig. 2). Although there was no significant correlation between total plasma levels of 1,25(OH)_2_D and 25(OH)D (Fig. 2A), total plasma levels of 24,25(OH)_2_D increased as a function of total plasma 25(OH)D levels (Fig. 2B).

**Figure 2.**
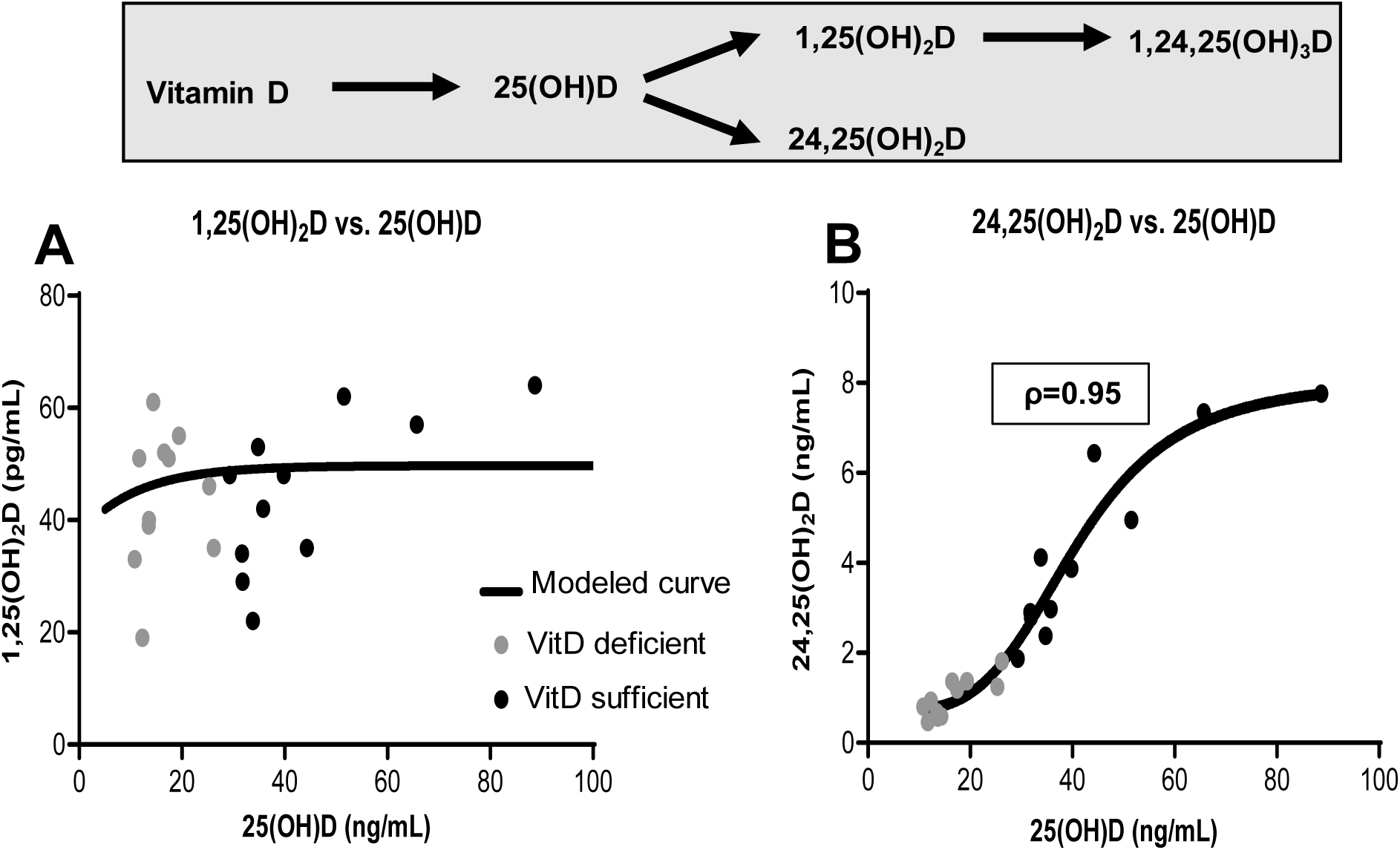
Total levels of 1,25(OH)_2_D and 24,25(OH)_2_D as a function of total levels of 25(OH)D. Panels A and B present pooled data for individual participants when they were VitD deficient (gray circles) and after they received VitD3 supplements (black circles). 1,25(OH)_2_D (panel A) and 24,25(OH)_2_D (panel B) are plotted as a function of total levels of 25(OH)D. The text box in panel B presents the Spearman correlation coefficient (ρ). The curves represent the equations derived from our mathematical model: equation (1) in Panel A and equation (7) in Panel B. Values of parameters for Equation (1): W_24_=8 ng/mL; L_24_=40 ng/mL; and α=0.1. Values for parameters for Equation (7): W_1,25_=40 ng/mL; L_1,25_=60 ng/mL; V_1_=40; K=60 ng/mL; α=0.1; and m_1,25_=1.

### A critical role for regulation of 24-hydroxylase activity in homeostatic regulation of 1,25(OH)_2_D

We considered possible mechanisms that might contribute to the observed sigmoid shape of the curve relating total 24,25(OH)_2_D levels to total 25(OH)D levels (Fig. 2B). If the 24-hydroxylase enzyme (CYP24A1) followed simple Michaelis-Menten kinetics, one might have predicted that the relationship between plasma levels of 24,25(OH)_2_D and 25(OH)D would have been described by a hyperbola (Fig. 3A, solid curve) rather than a sigmoid curve (Fig. 3A, dashed curve). The observed sigmoid curve approaches an asymptote at high levels of 25(OH)D – most likely a reflection of the saturability of the 24-hydroxylase enzyme (CYP24A1). What can explain the relative underproduction of 24,25(OH)_2_D at low levels of 25(OH)D as compared to what would have been predicted by simple Michaelis-Menten kinetics? As suggested by others (18, 23, 28), we hypothesized that expression of 24-hydroxylase enzyme is regulated, with low 24-hydroxylase activity occurring when 25(OH)D levels are low and higher 24-hydroxylase activity when 25(OH)D levels are high. We applied mathematical modeling (see Methods section) to provide quantitative estimates of the relationship between activity of the 24-hydroxylase enzyme and 25(OH)D levels (Fig. 3C). Our model suggests that 24-hydroxylase activity achieves a maximum when 25(OH)D levels exceed 50 ng/mL. In contrast, VitD deficiency is associated with suppression of 24-hydroxylase activity– achieving a minimum corresponding to ∼90% suppression when 25(OH)D levels are less than 10-20 ng/mL. Our mathematical model provides estimates of 24-hydroxylase activity as a function of 25(OH)D levels (Fig. 3C). The data points in Fig. 2A demonstrate substantial “scatter” around the modeled curve. We hypothesize that the ∼3-fold range of inter-individual variation in levels of VitD binding protein (VDBP) (29) may account for the observed 3.4-fold range of total levels of 1,25(OH)_2_D observed in this study, which in turn could account for the observed scatter for data points in the plot of total levels of 1,25(OH)_2_D versus total levels of 25(OH)D (Fig. 2A).

**Figure 3.**
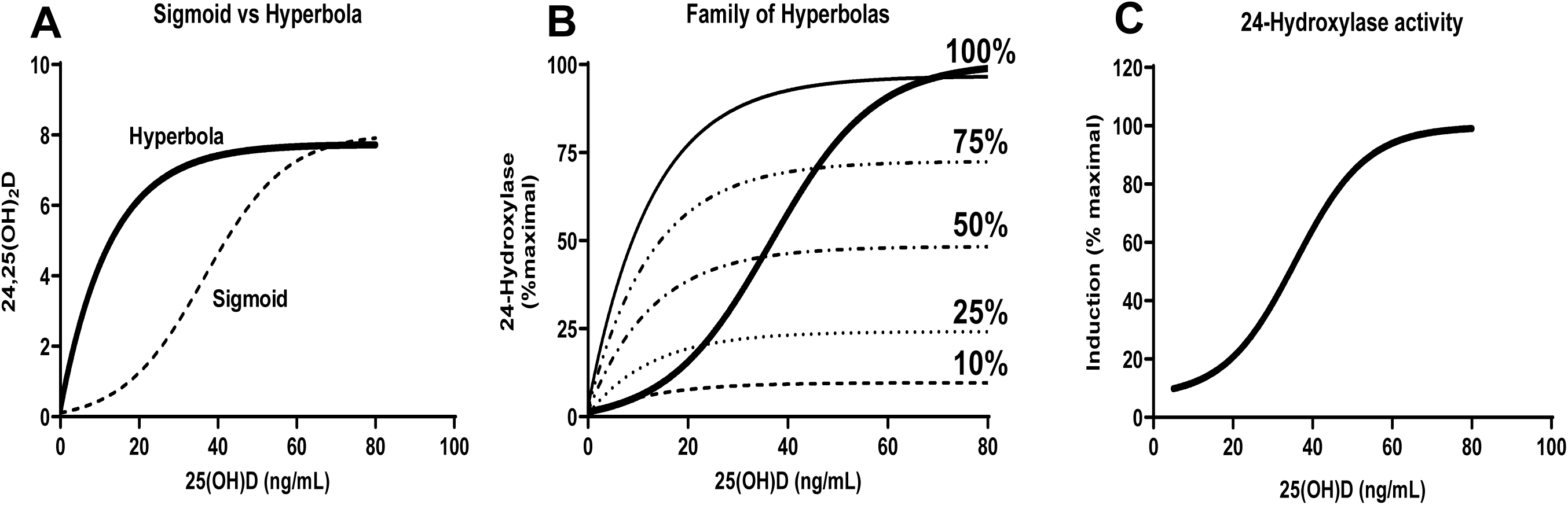
Assumptions underlying mathematical model: regulation of 24-hydroxylase activity in response to clinical VitD status. Panel A depicts curves corresponding to equations (1) and (2) [sigmoid and hyperbola, respectively]. Panel B depicts a family of hyperbolas [equation (2)] corresponding to varying degrees of induction from 10-100% of maximal induction of 24-hydroxylase activity. Panel C depicts equation (3), which is the fractional induction of 24-hydroxylase activity relative to the maximal level of induction as a function of 25(OH)D. Values of parameters for Equation (1): W_24_=8 ng/mL; L_24_=40 ng/mL; and α=.0.1. Values for parameters for Equation (2): V_24_=10 and K_24_=20 ng/mL. Values of parameters for Equation (3) m(24,25)=1; other parameters the same as for Equations (1) and (2).

Interestingly, the modeled curve predicts that levels of 1,25(OH)_2_D begin to decrease slightly for levels of 25(OH)D < ∼15-20 ng/mL. This “fall-off” may possibly be related to the fact that the model does not account for development of secondary hyperparathyroidism. If the model specified that low levels of 25(OH)D would trigger increased secretion of PTH and resultant increased 1α-hydroxylation of 25(OH)D, it is likely that the model would predict constant levels of 1,25(OH)_2_D for lower levels of 25(OH)D.

### Validating the model using the goodness of fit

Equation (1) provides a good fit to the observed data as shown by the graph plotting total levels of 24,25(OH)_2_D as a function of 25(OH)D (Fig. 2B). Goodness of fit is evidenced by the fact that the sum of the squared errors (SSE) is small relative to the total sum of the squares about the mean (SST): SSE/SST = 0.078 (Table 2).

**Table 2.** Goodness of fit for mathematical model equations. The Table includes metrics for the goodness of fit for the modeling equations for 1,25(OH)_2_D (Fig. 2A) and 24,25(OH)_2_D (Fig. 2B) as well as the goodness of fit for modeling of three VitD metabolite ratios (Figs. 4A, 5A, and 5B).

|  | 24,25(OH)D | 1,25(OH)<sub>2</sub>D | 1,25(OH)<sub>2</sub>D/<br>24,25(OH)<sub>2</sub>D | 25(OH)D/<br>1,25(OH)<sub>2</sub>D | 24,25(OH)<sub>2</sub>D/<br>25(OH)D |
| --- | --- | --- | --- | --- | --- |
| Sum of squares due to regression (SSR)<sup>a</sup> | 113 | 315 | 809 | 3.37 | 0.150 |
| Total sum of squares about mean (SST)<sup>b</sup> | 105 | 3341 | 849 | 3.22 | 0.151 |
| Sum of squared errors (SSE)<sup>c</sup> | 8.2 | 3483 | 412 | 1.66 | 0.079 |
| Ratio of SSE/SST | 0.078 | 1.04 | 0.49 | 0.52 | 0.52 |
| Ratio of SSR/SST | 1.08 | 0.094 | 0.95 | 1.05 | 0.995 |

<sup>a</sup> $SSR = \sum (\hat{x}_i - \bar{x})^2$
<sup>b</sup> $SST = \sum (x_i - \bar{x})^2$
<sup>c</sup> $SSE = \sum (x_i - \hat{x}_i)^2$
Where $x_i$ represents individual observed total levels of VitD metabolites, $\hat{x}_i$ represents individual predicted total levels of VitD metabolites, and $\bar{x}$ represents the mean of observed total levels of VitD metabolites.

Moreover, equation (7) correctly predicts that 1,25(OH)_2_D levels are not correlated with 25(OH)D levels (Fig. 2A). Indeed, the sum of the squared errors (SSE) is approximately equal to the total sum of the squares about the mean (SST): SSE/SST = 1.04 (Table 2). Furthermore, the value of the ratio of the sum of the squares due to regression (SSR) divided by SST = 0.094, suggesting that a horizontal line described by the following equation provides a good fit for the observed data: 1,25(OH)_2_D = 44.4 pg/mL (i.e., with 44.4 pg/mL being the mean level of 1,25(OH)_2_D).

### VitD metabolite ratios are better predictive of clinical VitD status

As noted in the literature, ratios of VitD metabolites are relatively independent of inter-individual variation in VDBP levels (24, 30, 31). For example, the following equations demonstrate that the ratio of [1,25(OH)_2_D]/[24,25(OH)_2_D] is independent of VDBP levels:

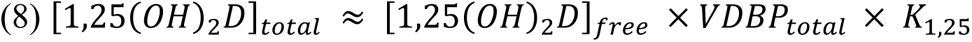

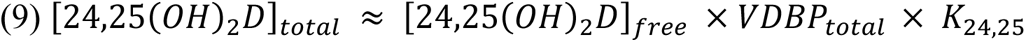

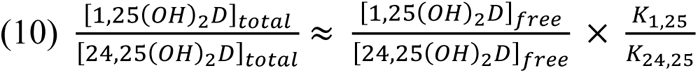

When 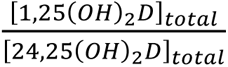 ratio is plotted as a function of total levels of 25(OH)D, this represents a graph of a VDBP-independent variable on the y-axis and a VDBP-dependent variable on the x-axis (Fig. 4A). In contrast, the graph of 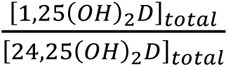 as a function of 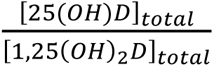 plots VDBP-independent variables on both axes (Figs. 4B). This approach minimized the contribution of inter-individual variation in VDBP levels, thereby minimizing “scatter” of points in the graph (Fig. 4B). When 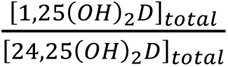 was plotted as a function of 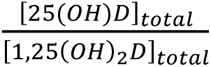, the absolute value of the Spearman correlation coefficient was quite high (ρ=-0.94; p=10^-10^) (Fig. 4B). In contrast, the absolute value of the Spearman correlation coefficient was lower (ρ=-0.83; p=2.2×10^-6^) when 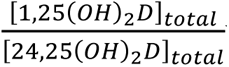 was plotted as a function of total levels of 25(OH)D (Fig. 4A).

**Figure 4.**
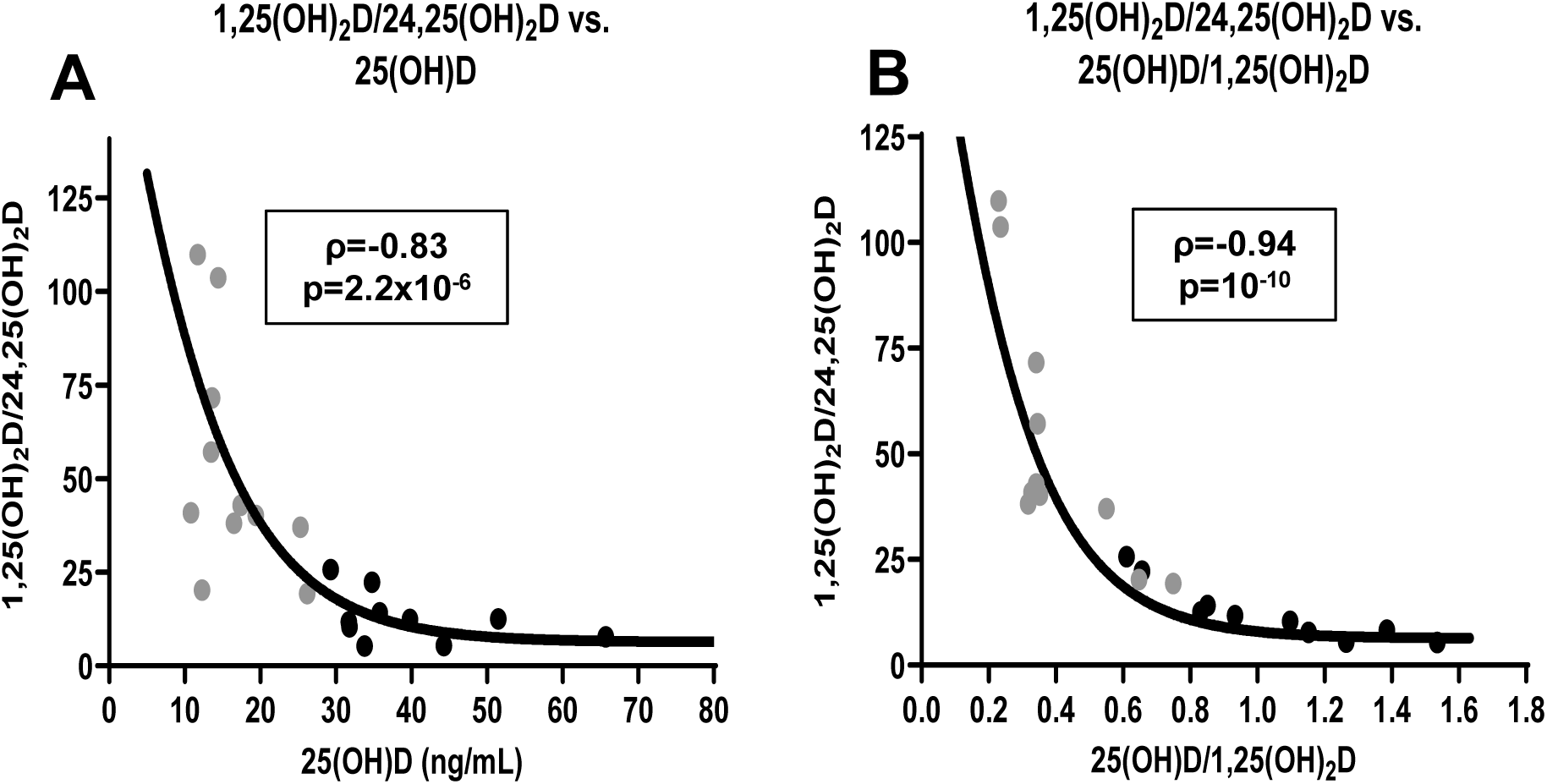
The 1,25(OH)_2_D/24,25(OH)D and 25(OH)D/1,25(OH)_2_D ratios reflect clinical VitD status. The graphs summarize data from individual participants when they were VitD deficient (gray circles) and after they received VitD3 supplements (black circles). Panel A depicts the 1,25(OH)_2_D/24,25(OH)_2_D ratio as a function of total 25(OH)D. The modeled curve was calculated as the ratio of modeled values for 1,25(OH)_2_D [equation (7)] divided by modeled values of 24,25(OH)_2_D [equation (1)]. Panel B depicts 1,25(OH)_2_D/24,25(OH)_2_D ratio as a function of 25(OH)D/1,25(OH)_2_D ratio. The fitted curves were calculated by GraphPad Prism software. Text boxes indicate Spearman correlation coefficients and p-values for the correlations.

### Modeling VitD Metabolite Ratios

Tang et al. (23) proposed that the ratio of 1,25(OH)_2_D/24,25(OH)_2_D provides an improved assessment of clinical VitD status with a ratio ≥51 suggesting “VitD deficiency” and a ratio of 35-50 suggesting “VitD insufficiency”. We applied our mathematical model to predict the shape of the curve expressing Tang’s VitD metabolite ratio (1,25(OH)_2_D/1,24(OH)_2_D) as a function of total levels of 25(OH)D. Our modeled curve (Fig. 4A) closely resembles the curve included in the publication by Tang et al. (23). Based on our modeled curve, the cutoffs proposed by Tang et al. correspond to total 25(OH)D levels of ∼17 ng/mL for “VitD deficiency” and ∼21 ng/mL for “VitD insufficiency”.

Furthermore, our mathematical model provides a good fit to the observed data as shown by the graph plotting total levels of 1,25(OH)_2_D/24,25(OH)_2_D as a function of 25(OH)D (Fig. 4A). Two other VitD metabolite ratios have been proposed in the literature: 1,25(OH)_2_D/25(OH)D (32) and 24,25(OH)_2_D/25(OH)D (24, 30, 31, 33, 34). Our mathematical model predicts that the 25(OH)D/1,25(OH)_2_D ratio [i.e., the reciprocal of the 1,25(OH)_2_D/25(OH)D ratio proposed in the literature (32)] is a near-linear function of 25(OH)D levels (Fig. 5A); the graph of the 24,25(OH)_2_D/25(OH)D ratio versus 25(OH)D is predicted to be a sigmoid curve (Fig. 5B). The modeled curves for all three VitD metabolite ratios provide good fits to observed data [SSE/SST ratios of 0.49-0.52 (Table 2)].

**Figure 5.**
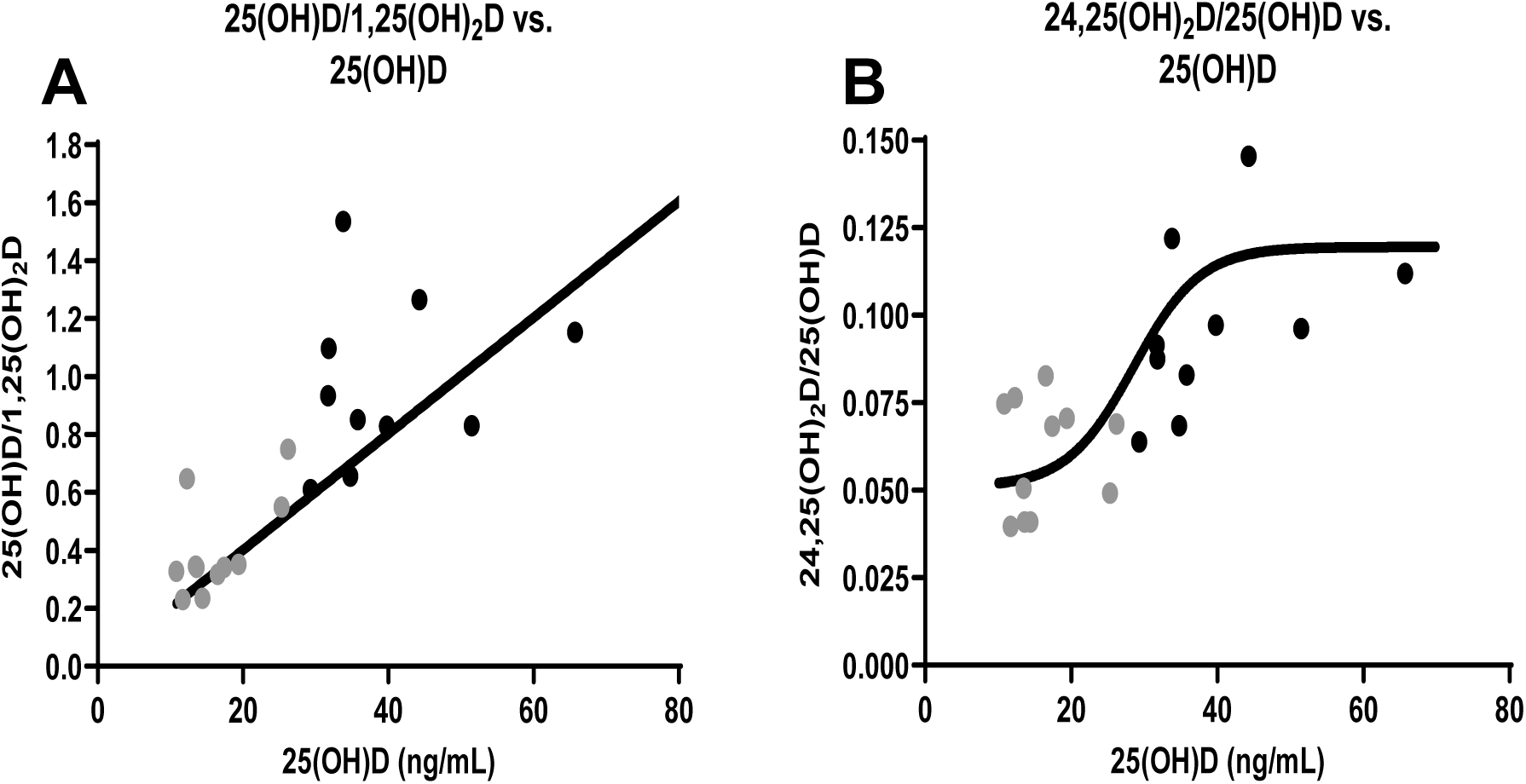
VitD metabolite ratios [25(OH)D/1,25(OH)_2_D and 24,25(OH)_2_D/25(OH)D] as a function of the total levels of 25(OH)D. The graphs summarize data from individual participants when they were VitD deficient (gray circles) and after they received VitD3 supplements (black circles). Panel A depicts the 25(OH)D/1,25(OH)_2_D ratio as a function of total 25(OH)D. The modeled curve was calculated as a linear regression using GraphPad Prism software. Panel B depicts 24,25(OH)_2_D/25(OH)D ratio as a function of 25(OH)D. The modeled curve was calculated as modeled values for 24,25(OH)_2_D [equation (1)] divided by assumed values of 25(OH)D.

### Correlation of relative 24-hydroxylase activity (h) with VitD metabolite ratios

We pooled our data before and after VitD3 supplementation for each of the three VitD metabolites. We first investigated the correlation of the calculated relative 24-hydroxylase activity (*h*) with total 25(OH)D levels, which gave a Spearman correlation coefficient of ρ=0.85 (p=7E-07) (Fig. 6A). We then evaluated correlations with three VitD metabolite ratios. As expected, relative 24-hydroxylase activity (*h*) was perfectly correlated with the 25(OH)D/1,25(OH)_2_D ratio (ρ=1) since this ratio was used in our model to account for the effect of variation in VDBP (Fig. 6B). While 24-hydroxylase activity (*h*) was highly correlated with the 1,25(OH)_2_D/24,25(OH)_2_D ratio (ρ^2^=0.94; p=3E-13) (Fig. 6C), *h* was less well correlated with the 24,25(OH)_2_D/25(OH) ratio (ρ^2^=0.67; p=3E-06) (Fig. 6D).

**Figure 6.**
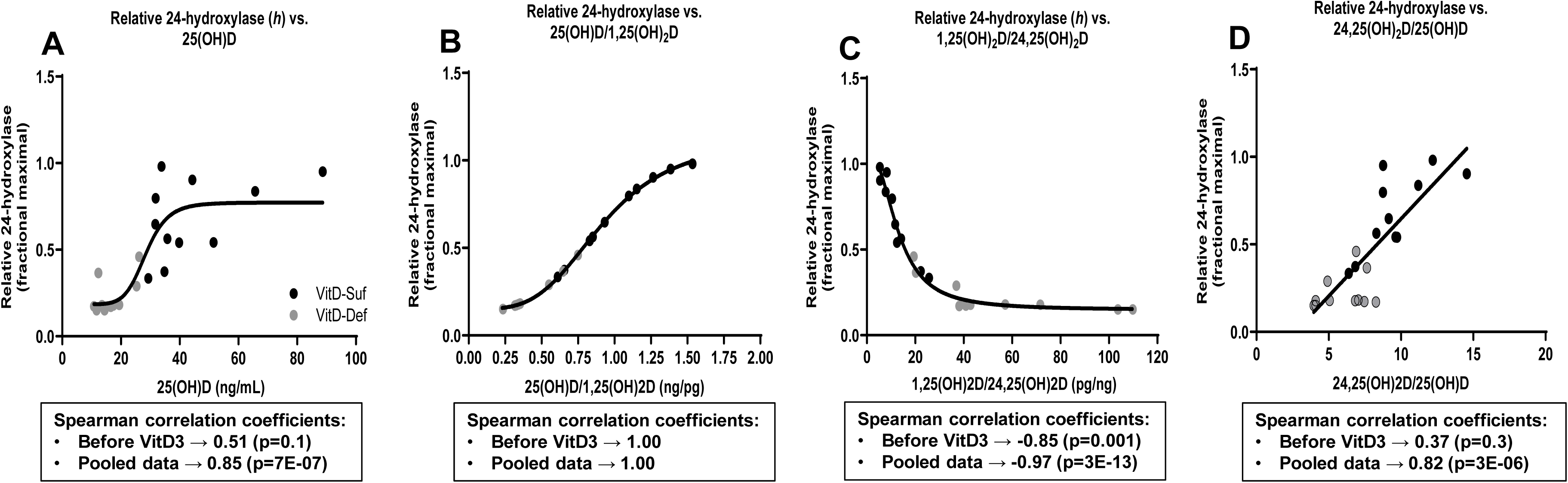
Correlation between relative 24-hydroxylase activity (*h*) and indices of VitD status. Panels A-D present pooled data when participants were VitD deficient (gray circles) and after they received VitD3 supplementation (black circles). The text boxes below the panels present two Spearman correlation coefficients and two p-values. The first bullet points correspond to the correlation between 24-hydroxylase and each of the VitD status indices at baseline before participants received VitD3 supplementation. The second bullet points correspond to the correlation between 24-hydroxylase and each of the VitD status indices based on the pooled data (both before and after VitD3 supplementation). The curves in the graphs were based on modeling of the pooled data.

### Correlation of baseline PTH with baseline indices of VitD status

Although baseline PTH levels were correlated with baseline total 25(OH)D levels before VitD3 supplementation (R^2^=0.08; p=5E-05), the variance in 25(OH)D accounted for only 8% of the variance in PTH. Furthermore, the slope of the regression line was positive whereas the known physiology predicts that the slope should be negative (Fig. 7A) with PTH levels being higher when 25(OH)D levels are lower. Moreover, baseline PTH levels were not significantly correlated with the baseline 1,25(OH)_2_D/24,25(OH)_2_D ratio (R^2^=0.008; p=0.63) (Fig. 7B). Even in participants who had received VitD3 supplementation, we observed a wide range for PTH levels (29-85 pg/ml) – suggesting that each participant has an individual setpoint for PTH levels.

**Figure 7.**
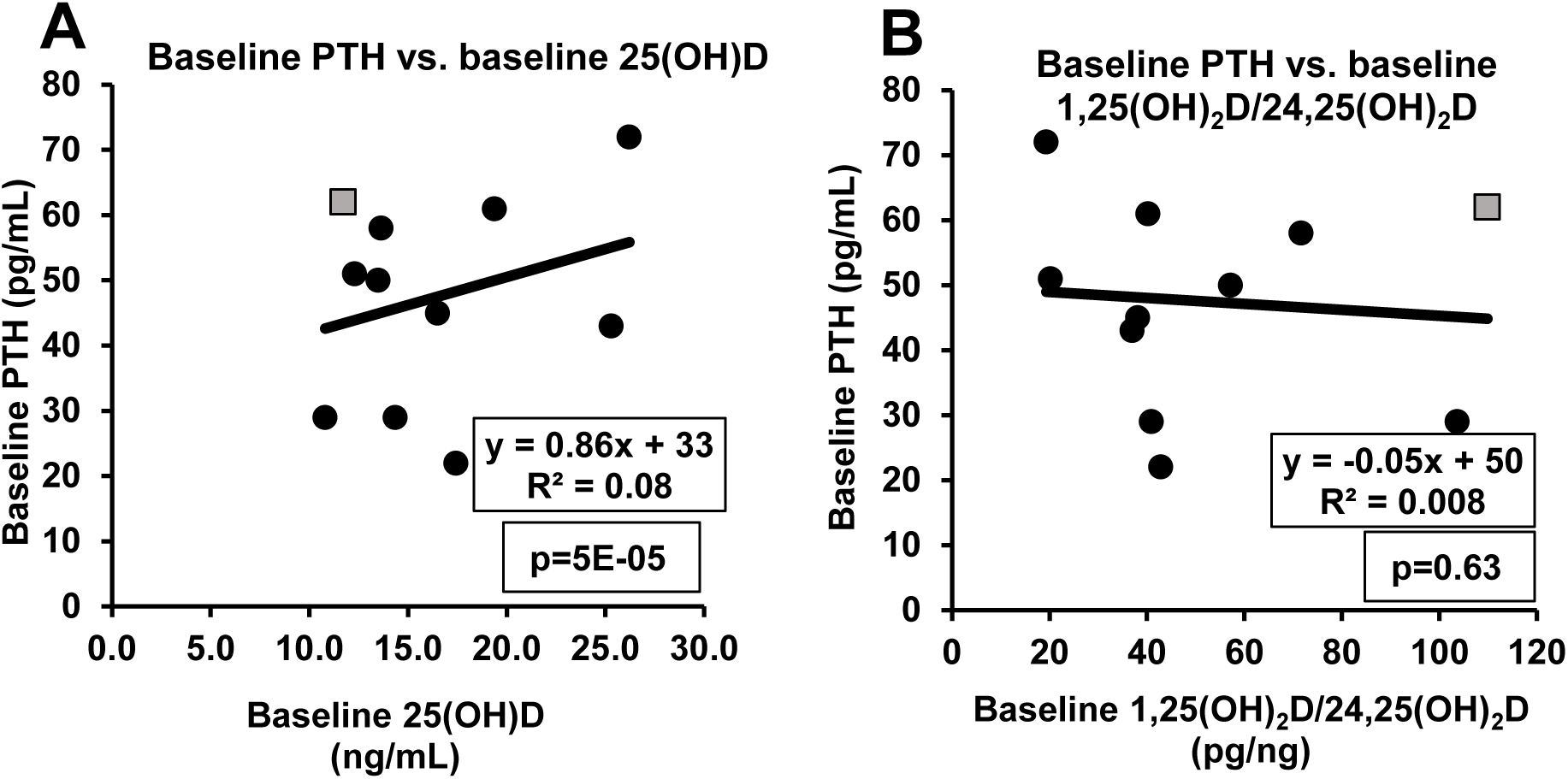
**Correlation between PTH and different indices of VitD status**. Panels A and B depict correlations of baseline PTH levels with baseline total 25(OH)D (panel A) or baseline 1,25(OH)_2_D/24,25(OH)_2_D ratio (panel B) before participants received VitD3 supplementation. One participant (designated by the gray square symbol) exhibited secondary hyperparathyroidism at baseline as evidenced by a 33% decrease in PTH levels in response to VitD3 supplementation. There was no statistically significant change in mean PTH levels in response to VitD3 supplementation (13). Interestingly, the participant with secondary hyperparathyroidism exhibited the highest baseline 1,25(OH)_2_D/24,25(OH)_2_D ratio among all the participants in our study (gray square in panel B).

## DISCUSSION

Because of the critical importance of Ca^+2^ in many biological processes, the body has evolved multiple homeostatic mechanisms to maintain free serum Ca^+2^ levels within a narrow physiological range. Since VitD plays a critical role in mediating intestinal absorption of dietary Ca^+2^, homeostatic mechanisms also maintain free 1,25(OH)_2_D within narrow physiological ranges. To maintain stable levels of 1,25(OH)_2_D when VitD availability is scarce, the body has two defense mechanisms: (1) to increase production of 1,25(OH)_2_D by increasing PTH-induced 1α-hydroxylation of 25(OH)D (12, 16, 17) and/or (2) to decrease the metabolic clearance of 1,25(OH)_2_D by suppressing 24-hydroxylase – the enzyme which degrades 1,25(OH)_2_D and 25(OH)D (14, 15). However, as noted in The Endocrine Society’s clinical practice guidelines, VitD deficiency-induced secondary hyperparathyroidism is associated with adverse effects on bone health: VitD-induced secondary hyperparathyroidism has an adverse impact on bone health by increasing “*osteoclastic activity thereby creating local foci of bone weakness, which causes a generalized decrease in bone mineral density, resulting in osteopenia and osteoporosis*” (17).

### Two lines of defense to maintain physiological levels of 1,25(OH)_2_D

Based on our study’s inclusion criteria, all research participants met The Endocrine Society’s criterion for VitD deficiency [i.e., screening 25(OH)D ≤ 20 ng/mL]. While total 25(OH)D and 24,25(OH)_2_D levels were increased significantly by VitD3 supplementation, total 1,25(OH)_2_D did not change. Moreover, levels of PTH were within normal limits at baseline and did not change when supplements were administered to correct the VitD deficiency. These data are consistent with published evidence suggesting that mild-moderate VitD deficiency may not trigger secondary hyperparathyroidism while secondary hyperparathyroidism is a characteristic feature of severe VitD deficiency [25(OH)D <10-12 ng/mL] (35–37).

The published literature contains many graphs displaying PTH levels as a function of total 25(OH)D (35, 38–41). In general, these graphs depict an increase in mean PTH levels as 25(OH)D levels decrease with an inflection point at total 25(OH)D levels in the range between 20-30 ng/mL (37). However, individual data points scatter widely above and below the mean values for PTH. If one focuses on mean data, one might conclude that all individuals are gradually becoming increasingly hyperparathyroid as total 25(OH)D decreases. However, detailed analysis of the data suggests an alternative interpretation. If one examines data points corresponding to total 25(OH)D levels in the range of 10-20 ng/mL, the vast majority of individual data points span the same wide range of PTH levels even as total 25(OH)D levels decrease (see for example Figs 2 and 3 from Mendes et al. (38)). However, the number of outliers (i.e., elevated PTH levels) begins to increase as total 25(OH)D levels decrease below ∼20 ng/mL. We interpret this observation as suggesting that there is considerable inter-individual variation in the setpoint for developing secondary hyperparathyroidism. When total 25(OH)D levels fall below ∼10 ng/mL, there is a substantial increase in both the mean level of PTH and the percentage of individuals with secondary hyperparathyroidism. We hypothesize that inter-individual variation in VDBP levels may be an important contributor. Individuals with high VDBP levels are predicted to have lower free 25(OH)D levels at any given level of total 25(OH)D. Thus, such individuals with high VDBP levels would be predicted to develop secondary hyperparathyroidism at higher levels of total 25(OH)D. Accordingly, we hypothesize that VDBP-independent indices of VitD status would better predict development of secondary hyperparathyroidism. Tang et al. (23) provided data suggesting that the 1,25(OH)_2_D/24,25(OH)_2_D ratio >100 is associated with an increased risk of elevated PTH level. Our data are consistent with the hypothesis that the 1,25(OH)_2_D/24,25(OH)_2_D ratio may be better correlated with secondary hyperparathyroidism as compared to other indices of VitD status such as the total 25(OH)D level.

The absence of secondary hyperparathyroidism in our participants with total 25(OH)D in the range between 11 to 19 ng/mL inspired us to investigate what other homeostatic mechanisms maintained 1,25(OH)_2_D in the normal range. Analysis of our data demonstrated that the correlation between 25(OH)D and 24,25(OH)_2_D levels was defined by a sigmoid curve rather than a hyperbola. Based on this observation, we hypothesized that 24-hydroxylase expression is regulated, and we designed a mathematical model to test our hypothesis. Our model demonstrates that 24-hydroxylase activity achieves a minimum (∼90% suppression) when 25(OH)D levels <10-20 ng/mL. Thus, in mild-moderate VitD deficiency, the body suppresses 24-hydroxylation to preserve 1,25(OH)_2_D from degradation and this mechanism provides the first line of defense against VitD deficiency. Interestingly, viewed from a different perspective, high levels of 25(OH)D are associated with increased 24-hydroxylase activity, which provides a major defense against VitD toxicity (26, 27). In the absence of secondary hyperparathyroidism, it is possible that mild-moderate VitD deficiency may not represent an absolute requirement for treatment with VitD supplements if the treatment objective is to promote bone health. On the other hand, recent publications have suggested that 24,25(OH)_2_D exerts a direct effect to promote healing of bone fractures (42–44). In other words, whether or not VitD deficiency increases the risk of bone fracture (45, 46), it has been proposed that adequate levels of 24,25(OH)_2_D may promote healing of bone fractures.

Considerable controversy exists with respect to the overall health impact of VitD status and VitD supplementation in the general population (45–48). Published literature demonstrates that severe VitD deficiency triggers secondary hyperparathyroidism, which in turn induces 1α-hydroxylation – thereby increasing the rate of 1,25(OH)_2_D biosynthesis (16, 35). Because secondary hyperparathyroidism is not triggered unless VitD deficiency becomes severe [e.g., 25(OH)D <10-12 ng/mL] or if 1α-hydroxylation is impaired as in chronic kidney disease, this mechanism provides the second line of defense against VitD deficiency. Secondary hyperparathyroidism is associated with adverse effects on bone health. This provides a strong rationale for treatment with VitD supplements. We propose two diagnostic terms to reflect clinical differences between these two scenarios. “Sub-clinical VitD deficiency” describes a clinical condition when only the first line of defense (suppression of 24-hydroxylase) has been triggered. In this manuscript, we have used the term “mild-moderate” VitD deficiency as a synonym for the concept of sub-clinical VitD deficiency (i.e., suppression of 24-hydroxylase in the absence of secondary hyperparathyroidism). “Overt VitD deficiency” describes a clinical condition when both lines of defense are triggered (i.e., suppression of 24-hydroxylase plus secondary hyperparathyroidism). This is consistent with conclusions of a scientific workshop sponsored by the National Kidney Foundation in 2018, which suggested that in patients with chronic kidney disease, 25(OH)D levels of <15 ng/mL require VitD treatment and in the range of 15-20 ng/mL, require treatment only if there is evidence of secondary hyperparathyroidism (22).

Which molecular mechanism(s) mediate homeostatic regulation of 24-hydroxylase activity? Expression of the *CYP24A1* gene (the gene encoding 24-hydroxylase enzyme) is reported to be increased by 1,25(OH)_2_D and FGF23 but decreased by PTH (49). However, mean levels of total 1,25(OH)_2_D, FGF23, and PTH did not change in response to VitD3 supplementation in our clinical trial (13). Taken at face value, these observations suggest that regulation of 24-hydroxylase activity may be mediated by factors other than levels of 1,25(OH)_2_D, FGF23, or PTH. Nevertheless, it remains possible that regulation of 24-hydroxylase activity is sensitive to extremely small changes in levels of 1,25(OH)_2_D, FGF23, or PTH, which were not captured in our study. Furthermore, all three hormones exhibit circadian variation (50–52). Since we obtained blood samples at only one time of day, we cannot exclude the possibility that the relevant biomarker(s) might require measurements of circulating levels integrated over a 24-hour period. In any case, additional research will be required to answer this interesting question.

### Implications for clinical diagnosis of VitD deficiency

Although the Endocrine Society’s clinical practice guidelines recommend a “one-size-fits-all” approach based on measurements of total levels of 25(OH)D (17), this approach does not account for the substantial inter-individual variation in levels of VDBP that limits the applicability of a one-size-fits-all approach. Whereas the usual clinical assays primarily reflect protein-bound levels of VitD metabolites, it is the free levels (i.e., not bound to protein) that are biologically relevant (53). This provides a strong rationale for a *precision diagnostics* approach wherein diagnostic criteria for VitD deficiency are adjusted for each individual based on that person’s circulating level of VDBP. Other factors may also be relevant: for example, individual setpoints for PTH secretion and an individual’s sensitivity to biological actions of PTH (54). These considerations raise challenging questions about how best to implement a precision diagnostics approach to diagnose VitD deficiency and secondary hyperparathyroidism. Others have suggested the potential value of using a tailored approach to assess VitD status and to guide decisions about the need for VitD supplementation (55). In clinical endocrinology practice, deficiency states are frequently diagnosed by documenting the activation of compensatory homeostatic mechanisms. For example, hypothyroidism is diagnosed by documenting elevated levels of TSH. Conversely, hyperthyroidism is diagnosed by documenting suppression of TSH levels (20). By basing diagnoses on whether the body’s homeostatic responses have been triggered, physicians implicitly rely on Cannon’s concept of the “*wisdom of the body”* to diagnose hormonal status. Following these precedents, one might base the diagnosis of VitD deficiency on documenting the activation of the relevant homeostatic mechanisms – e.g., suppression of 24-hydroxylase activity. However, it is challenging to base the diagnosis solely on measuring total 24,25(OH)_2_D levels because of the confounding influence of inter-individual variation in VDBP levels. The observed threefold inter-individual variation in levels of VDBP (29) is predicted to cause a three-fold inter-individual variation in total levels of 24,25(OH)_2_D even in individuals with identical levels of free 24,25(OH)_2_D. Fortunately, VitD metabolite ratios are known to be independent of VDBP levels (24). Tang et al. (23) have advocated the use of the 1,25(OH)_2_D/24,25(OH)_2_D ratio for this purpose. As confirmed by our mathematical model (Fig. 4A), VitD deficiency is associated with a dramatic increase in the 1,25(OH)_2_D/24,25(OH)_2_D ratio as a consequence of the marked suppression of 24-hydroxylase activity induced by VitD deficiency. By providing an index of the level of 24-hydroxylase activity, the 1,25(OH)_2_D/24,25(OH)_2_D ratio provides a highly sensitive index of clinical VitD status. Furthermore, because of the extremely high correlation between the 1,25(OH)_2_D/24,25(OH)_2_D ratio and the 25(OH)D/1,25(OH)_2_D ratio (Fig. 4B), it seems likely that the 25(OH)D/1,25(OH)_2_D ratio might also provide a useful clinical assessment of VitD status. This latter ratio is based on two widely available assays [25(OH)D and 1,25(OH)_2_D] whereas the 1,25(OH)_2_D/24,25(OH)_2_D ratio requires an assay for 24,25(OH)_2_D, which is currently less widely available and more expensive. Different VitD metabolite ratios have been suggested for assessment of VitD status and some studies have investigated their predictive value for hard clinical endpoints e.g., risk of bone fracture (31, 33). The 24,25(OH)_2_D/25(OH)D ratio has gained attention as a clinically informative substitute for total plasma 25(OH)D for assessment of VitD status. Furthermore, the scientific workshop sponsored by the National Kidney Foundation proposed that the 24,25(OH)_2_D/25(OH)D ratio might be particularly useful in patients with CKD as a surrogate for activity of 24-hydroxylase, as in this patient population 1α-hydroxylation of 25(OH)D is impaired (22). However, our data suggest that the 1,25(OH)_2_D/24,25(OH)_2_D ratio may have superior predictive value as compared to the 24,25(OH)_2_D/25(OH)D ratio.

### Interindividual variation in PTH levels

Even in VitD-replete participants being treated with VitD3 supplements, we observed a wide range of PTH levels (29-85pg/mL). Furthermore, mean levels of PTH were not significantly affected by administration of VitD3 supplements in our study (47.5±4.8 pg/mL before and 48.4±5.1 pg/mL after VitD3 supplementation). As suggested by others (56), interindividual variation in PTH levels may reflect variation in the setpoint for PTH secretion. Variation in dietary calcium intake may also affect PTH levels (57). These considerations create major challenges in how to interpret an individual measurement of PTH. We, therefore, assessed the response of PTH to VitD3 supplementation as an indication of whether a participant exhibited secondary hyperparathyroidism due to VitD deficiency. While one participant exhibited a ∼34% decrease in PTH level in response to VitD3 supplementation, the other participants did not exhibit significant suppression of PTH levels in response to VitD3 supplementation. Consistent with the conclusions of Tang et al. (23), this patient with secondary hyperparathyroidism exhibited the highest value of the 1,25(OH)_2_D/24,25(OH)_2_D ratio (110 pg/ng) observed in our study. Furthermore, the 1,25(OH)_2_D/24,25(OH)_2_D ratio was highly correlated (ρ^2^=-0.94; p=3E-13) with relative 24-hydroxylase activity as calculated by our mathematical model – accounting for 94% of the variance in relative 24-hydroxylase activity. Other indices were less well correlated: 24,25(OH)_2_D/25(OH)D (ρ^2^=0.67; p=3E-06) and total 25(OH)D (ρ^2^=0.72; p=7E-07).

### Study design: strengths and limitations

The study was designed as a crossover study to enable paired statistical analyses comparing data obtained before versus after administration of VitD3 supplements. To minimize the impact of circadian rhythms for plasma 1,25(OH)_2_D, PTH, and FGF23 (50–52), all blood samples were obtained at the same time of day (∼7:00 am). Using LC-MS/MS-based assays of VitD metabolites (Quest Diagnostics and Heartland Assays), we did not detect metabolites of VitD2 in any of our participants. The absence of VitD2 metabolites simplified data interpretation by avoiding potential issues related to differences between VitD2 and VitD3 (58). Data on levels of 25(OH)D and 24,25(OH)_2_D were provided by Heartland Assays – a company that specializes in applying liquid chromatography and mass spectroscopy (LC-MS/MS) to conduct scientifically rigorous and reproducible assays of VitD metabolites. Our novel mathematical model provided quantitative estimates of *in vivo* 24-hydroxylase activity and represents another strength of the study. Furthermore, while previous publications provide data on one or another ratio of VitD metabolites (22, 24, 30, 31), our analysis compares the predictive value of three VitD metabolite ratios: 1,25(OH)_2_D/24,25(OH)_2_D; 25(OH)D/1,25(OH)_2_D; and 24,25(OH)_2_D/25(OH)D.

In addition to these strengths of our study design, the study had important limitations. Although our original study design proposed to study 25 participants, the impact of the COVID-19 pandemic necessitated a decrease in the sample size to 11 participants. Limitations associated with the relatively small sample size are partially mitigated by our efforts to assure scientific rigor and reproducibility of the primary data – e.g., the enhanced statistical power provided by conducting paired statistical comparisons. Accordingly, many of our analyses [e.g., VitD-induced changes in 25(OH)D and 24,25(OH)_2_D] demonstrated high levels of statistical significance. Other analyses [e.g., comparisons for 1,25(OH)_2_D and PTH] demonstrated essentially identical mean values before and after VitD3 supplementation. Based on clinical guidelines of The Endocrine Society, all of our participants exhibited some degree of VitD deficiency. However, only one participant exhibited more severe VitD deficiency accompanied by secondary hyperparathyroidism. Additional research will be required to better understand VitD homeostatic mechanisms in patients with severe VitD deficiency.

We followed The Endocrine Society’s clinical guidelines recommending a loading dose of VitD3 (i.e., 50,000 IU/week for 4 weeks) with a modification by increasing the dose in patient with BMI ≥30 kg/m^2^). Some authorities have raised questions about the impact of high dose boluses of VitD that transiently raise VitD concentrations to supraphysiological levels (59). However, we minimized this concern by maintaining participants on maintenance doses of VitD3 (1000-2000 IU/day depending on BMI) for a minimum of six weeks. This allowed for a re-equilibration to a more physiological level of VitD.

Finally, this study was designed as an outpatient study with participants free to eat *ad libitum*. Day-to-day variation in dietary intake of calcium and phosphate may have increased variation in measurements of various endpoints – e.g., PTH or FGF23. Furthermore, since our study design did not include data on dietary intake, our analysis could not investigate dietary factors as covariates. We have offered some tentative hypotheses related to the necessity of VitD supplements in overt VitD deficiency as compared to the possible optional nature of VitD supplements in sub-clinical VitD deficiency. However, we acknowledge that the present study does not provide sufficient data to draw rigorous evidence-based conclusions with respect to treatment recommendations. Our small pilot study is based on biomarker data rather than hard clinical outcomes. Future research will be required to address these important clinical questions. Nevertheless, we believe it is valuable to offer hypotheses based on implications derived from underlying physiology and pathophysiology.

## Acknowledgements

The authors gratefully acknowledge research funding provided by NIDDK and the NIH Office of Dietary Supplements: R01DK118942, R01DK118942-02S1, and T32DK098107. We are also grateful to the staff at University of Maryland’s Amish Research Clinic and to the members of the Old Order Amish Community in Lancaster, PA who participated in this clinical trial.

## Data Availability

Primary data will be made available upon request to qualified academic investigators for research purposes under a Data Transfer Agreement to protect research participants’ confidential information.

## Trial registration

NCT02891954 (clinicaltrials.gov)

